# Translation and Cultural Adaptation of the MYCaW® Questionnaire into German: The iSWOP Study Protocol

**DOI:** 10.1101/2025.03.17.25324105

**Authors:** Anja Thronicke, Lisa Schille, Katja Adie, Christian Junghanss, Shiao Li Oei, Sophia K. Johnson, Juliane Roos, Friedemann Schad

## Abstract

**Introduction:** The growing population of cancer survivors faces persistent physical and emotional challenges that significantly impact health-related quality of life (HRQL). To address these multifaceted needs, robust and culturally adapted patient-reported outcome measures are essential for understanding and improving survivors’ subjective experiences. This protocol outlines the systematic translation and cultural adaptation of the Measure Yourself Concerns and Wellbeing (MYCaW®) questionnaire into German. The MYCaW® questionnaire, a patient-reported outcome measure, is designed to capture individualized concerns and assess overall well-being, particularly in cancer care settings. By adhering to common guidelines, this research will provide a tool for assessing individualized concerns and patient needs among German-speaking cancer patients.

**Methods and analysis:** Following International Society for Pharmacoeconomics and Outcomes Research (ISPOR) guidelines, this study will employ a structured methodology involving forward and backward translation, expert review, patient review process, and preliminary validation to ensure linguistic and cultural equivalence. The study’s strengths (systematic methodology, patient-centered approach, expert oversight, clinical utility) and limitations (preliminary validation sample size, potential bias in translation) provide a balanced view of the study’s rigorous design and areas where further research or refinement could improve its reliability and generalizability.

**Ethics and disseminations:** Ethics committee of the the Medical Association Berlin (Ärztekammer Berlin) (reference number Eth-27/10) gave ethical approval for this work. The findings will be presented at scientific conferences and submitted for publication in peer-reviewed journals.

**Trial registration number:** The study was registered at the German Register for Clinical Trials under DRKS00013335 on 27/11/2017.

## Introduction

A growing number of people are living with cancer or have survived it, making the well-being of these individuals an important area for research [1]. In addition to the advances in early detection and the increasing efficacy of oncology therapies, understanding and improving the health-related quality of life (HRQL) of cancer patients has gained increasing attention. Many cancer patients face persistent physical and emotional challenges long after completing primary therapy. Long-term and late effects of cancer treatments—such as chronic pain, cognitive dysfunction, and severe fatigue—can significantly impact HRQL [2,3]. These impairments often necessitate comprehensive strategies to address the multifaceted needs of oncology patients.

The assessment of HRQL relies on validated patient-reported outcome measures (PROMs). These tools provide invaluable insights into patients’ subjective experiences, aiding clinicians and researchers in identifying specific areas requiring intervention. One such tool, the Measure Yourself Concerns and Wellbeing (MYCaW®) questionnaire, is recognized for its ability to capture individualized concerns and overall well-being [4]. MYCaW® employs a patient-centred approach, allowing participants to report their primary concerns and assess their well-being, making it particularly useful in integrative oncology settings. The MYCaW® questionnaire was first introduced in 2006 by Dr. Charlotte Paterson [4], a postdoctoral researcher at the University of Bristol, with support from the MRC Health Services Research Collaboration. Earlier work on MYCaW® at the Bristol Cancer Help Centre was recognized for its impact, earning a place on the shortlist and winning the Healthcare and Medical Research category at the 2003 Charity Awards. In 2020 Dr. Polley and Dr. Seers took over responsibility for MYCaW® and MYMOP® leading to the foundation of Meaningful Measures Ltd [16, 17, 18] to manage the licensing of the questionnaire [19] while providing tailored guidance on capturing person-centered outcomes. The cross-cultural adaptation and validation of PROMs are essential to ensure their relevance and applicability across different languages and cultural contexts [5]. Translation is a critical first step in this process, requiring not only linguistic precision but also cultural sensitivity to maintain the questionnaire’s conceptual equivalence and reliability. To guide this process, the widely recognized framework by Beaton et al. [5] outlines a systematic multi-step methodology involving forward and backward translations, expert review, and patient review processes to ensure conceptual equivalence between the original and adapted versions. Additionally, the International Society for Pharmacoeconomics and Outcomes Research (ISPOR) principles for good practices in translation and cultural adaptation emphasize maintaining linguistic precision and cultural sensitivity [6], while the Consensus-based Standards for the selection of health status Measurement Instruments (COSMIN) guidelines provide recommendations for assessing the measurement properties of translated instruments [7], such as validity, reliability, and cross-cultural equivalence. Together, these frameworks ensure that adapted tools are both culturally relevant and psychometrically sound.

This study protocol outlines the process of translating the MYCaW® questionnaire into German and aims to provide a detailed framework for its cultural adaptation. By achieving a validated German version, this research aims to facilitate its use among German-speaking cancer patients, enabling a deeper understanding of their self-reported individualized concerns and needs.

The primary objective of this study is to describe the methodology employed in the translation and adaptation of the MYCaW®® questionnaire into German. Ultimately, the study aims to contribute to the broader field of patient-centred oncology care by enabling more accurate and culturally appropriate assessments of HRQL in German-speaking settings.

## Methods

### 1. Study design

This study will follow the ISPOR guidelines for the translation and cultural adaptation of patient-reported outcome measures [6] and the COSMIN guidelines for assessing cross-cultural validity and measurement invariance [7], see supplementary file 1. The process will include four key phases: preparation, translation, patient review, and validation.

### 2. Study setting

This study is conducted by the Research Institute Havelhöhe (Forschungsinstitut Havelhöhe, FIH) at the hospital Gemeinschaftskrankenhaus Havelhöhe in Berlin (GKHB) within the Network Oncology (NO) [8].

### 3. Translation Process

In figure 1 the flowchart illustrating the systematic process of the questionnaire translation into German language following the ISPOR guidelines. The steps ensure linguistic and conceptual equivalence between the original and translated versions, maintaining validity and reliability.

**Figure 1.**
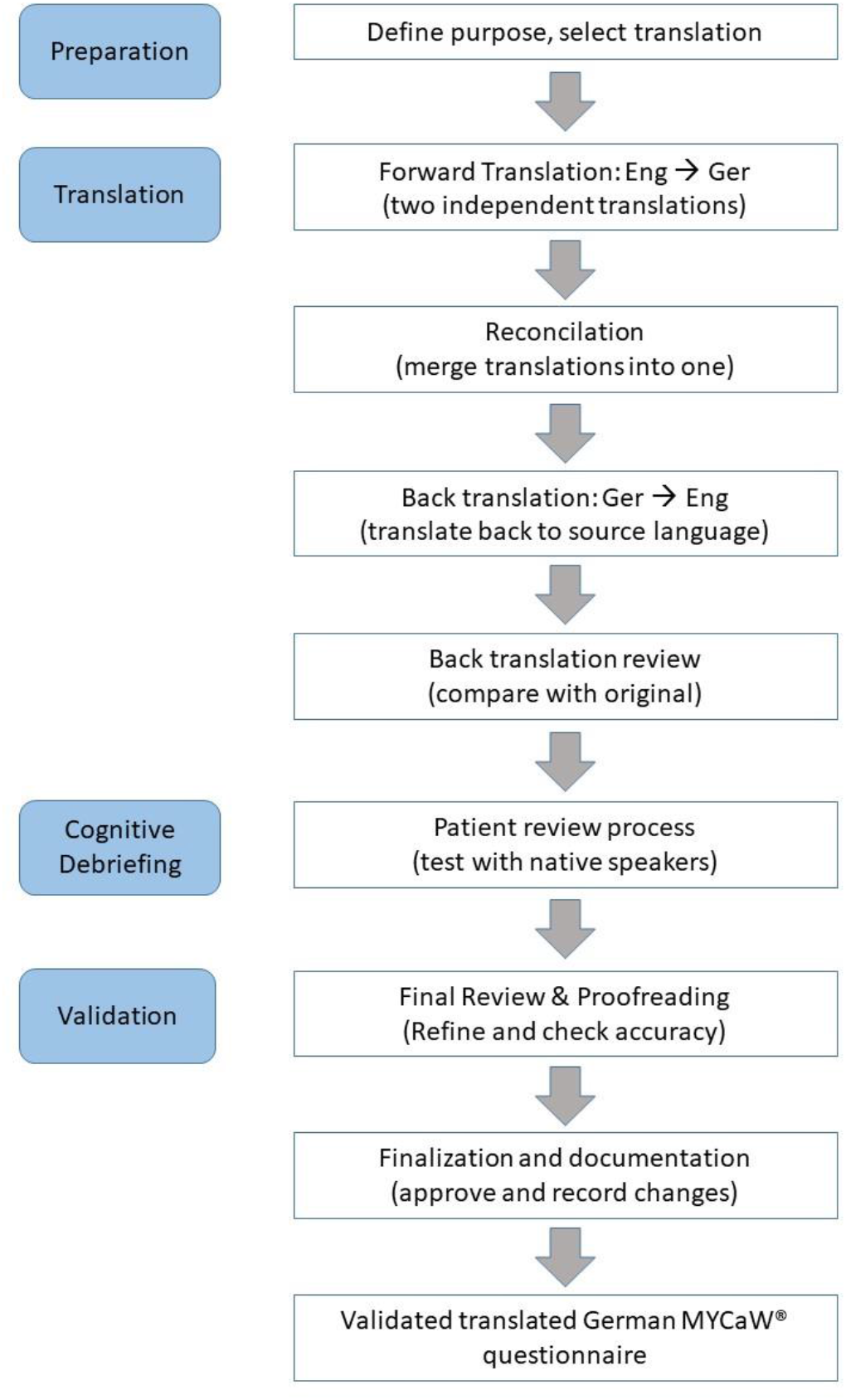
Flow chart of the translation process according to ISPOR guidelines; Ger, German language; Eng, English language; ISPOR, International Society for Pharmacoeconomics and Outcomes Research

#### 3.1 Forward & Backward Translation, Expert Committee Review, Patient Review, and Preliminary Validation

Two independent bilingual translators (native German speakers fluent in English) will produce two German versions of the MYCaW® questionnaire, see figure 1. Translators will focus on maintaining conceptual equivalence rather than literal translation, considering cultural nuances and medical terminology. A reconciliation meeting with both translators and a third expert will combine the two forward translations into a single German draft. Two different independent translators (native English speakers fluent in German) will back-translate the reconciled German version into English. The backward translations will be compared with the original MYCaW® questionnaire to identify discrepancies, see figure 1. A panel of experts, including oncologists, psychologists, and patient representatives, will review all versions to ensure linguistic and cultural accuracy. The panel will finalize a pre-test version of the German MYCaW® questionnaire. The pre-test version will be administered to 15 cancer patients in Germany to evaluate its clarity, cultural relevance, and ease of understanding. Semi-structured interviews will be conducted to gather participants’ feedback on specific items, see figure 1. Table 1 presents the timeline of the MYCaW® translation and validation process. Key milestones and projected completion dates are highlighted, with further details available in figure 2 (Gantt chart). The finalized German version (see supplementary material) will undergo preliminary testing with a larger sample to assess reliability and validity, see table 1. The psychometric properties to be evaluated include content validity, internal consistency, and construct validity. These psychometric evaluations are critical to ensuring that the translated MYCaW® questionnaire is both reliable and valid for assessing health-related quality of life among German-speaking oncological patients.

**Table 1.** Timeline of the study

| **2023 - Translation Process Completion (Phase 1)** |  |
| --- | --- |
| Q4 2023 –<br>Q2 2024 | Finalize the German MYCaW® version after translation, reconciliation, and expert review |
| **2024 – Patient Review Process and Preliminary Validation (Phase 2 - 3)** |  |
| Q2 2024 –<br>Q3 2024 | Patient review process<br>Test pre-final version with 15 patients, conduct interviews, refine based on feedback |
| Q3 2024<br>Q4 2024 | Finalize the German version, begin validation with a larger sample of participants, assess psychometric properties (content validity, internal consistency, construct validity) |
| **2025 - Data Analysis, Final Testing, and Study Conclusion (Phase 4 -6)** |  |
| Q4 2024 –<br>Q3 2025 | Continue validation, finalize data collection<br>Complete recruitment, conduct final psychometric analysis, review preliminary results |
| Q3 2025 –<br>Q4 2025 | Conclude study, finalize participant follow-up<br>Publish findings, present results, and complete study |
| **December 2025 - Study officially ends** |  |

**Figure 2.**
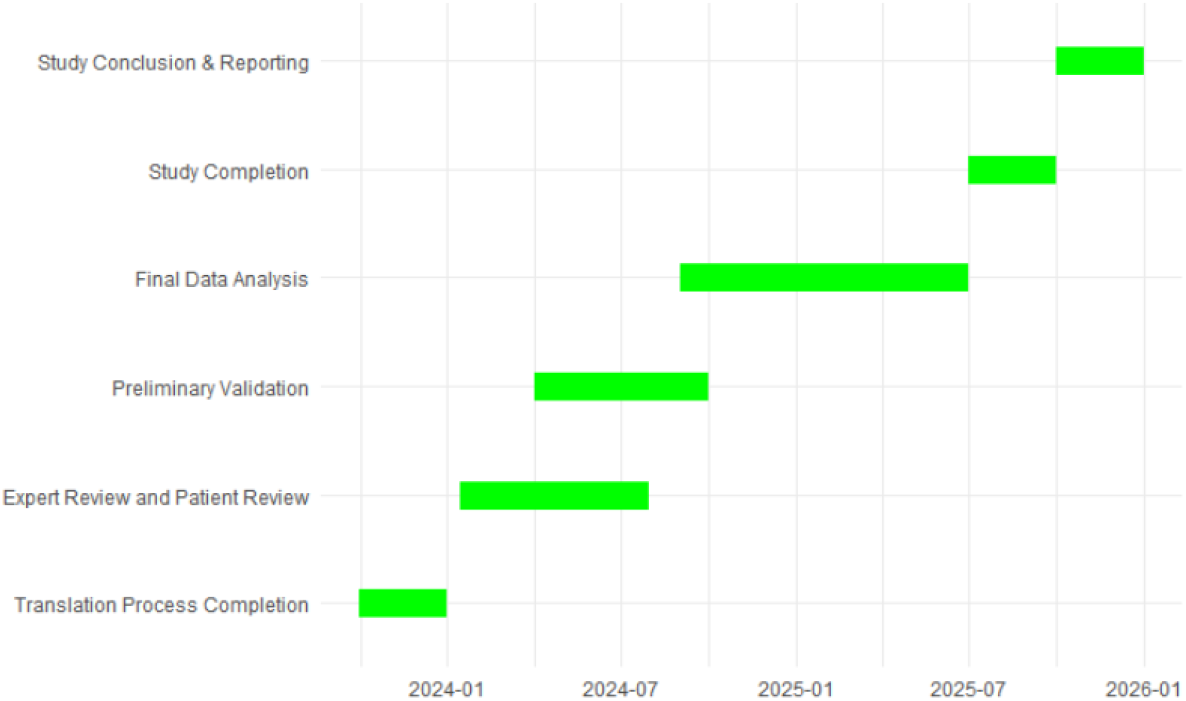
Milestone plan of the iSWOP study.

#### 3.2 Content Validity, Internal Consistency and Construct Validity

Content validity will be assessed through patient feedback, which helps ensure that the items included in the questionnaire are relevant, comprehensive, and representative of the experiences of cancer patients. This process typically involves gathering input from a sample of patients through methods such as interviews or surveys. By comparing the content of the MYCaW® questionnaire with the concerns and well-being issues reported by participants, researchers can confirm that the tool effectively captures the key aspects of HRQL that are important to the target population. Cronbach’s alpha, with values ranging from 0 to 1, with higher values indicating greater reliability, will be used to measure internal consistency. A commonly accepted threshold for good internal consistency is a Cronbach’s alpha of 0.7 or higher. This will be calculated for each subscale of the MYCaW® questionnaire to ensure that the items within each dimension of HRQL are consistent and reliable in measuring the same underlying construct. To assess construct validity, the MYCaW® questionnaire will be compared with other well-established HRQL measures to examine whether the MYCaW® questionnaire captures similar constructs and whether changes in MYCaW® scores align with changes in other established measures. This approach helps ensure that the MYCaW® questionnaire is measuring HRQL in a manner consistent with other validated tools. Figure 2 presents the Gantt chart outlining the timeline of the MYCaW® translation process. Key milestones and projected completion dates are highlighted, with further details available in Table 1.

### 4 Strengths and limitations of this study

The strength of this study is its systematic methodology as it follows internationally recognized guidelines (ISPOR and COSMIN) to ensure a rigorous translation and cultural adaptation process. By incorporating cognitive debriefing and semi-structured interviews, the study ensures that the MYCaW® questionnaire is linguistically accurate and culturally relevant and meaningful to German-speaking cancer patients. The validated German MYCaW® questionnaire will provide healthcare professionals with a tool to better understand and address individualized concerns and needs. A limitation of the study could be the sample size, which may not capture the full diversity of oncological patients, although it is adequate for initial psychometric testing. Despite efforts to minimize discrepancies, inherent biases in forward and backward translation could affect the conceptual equivalence of the translated version.

## Discussion

This study aims to ensure the linguistic and cultural appropriateness of the German MYCaW® questionnaire. The methodology outlined here can serve as a blueprint for similar cross-cultural adaptation studies. The MYCaW® questionnaire has been widely utilized in various research settings to assess patient-reported concerns and overall well-being. Its flexible and patient-centered design allows individuals to express their specific concerns, making it particularly suitable for integrative oncology and supportive care settings [4].

Several studies have employed MYCaW® to explore the impact of interventions, gauge patient priorities, and assess changes in well-being over time [9]. Previous research on MYCaW® has demonstrated its utility in oncology care by capturing nuanced, patient-specific concerns that standardized questionnaires may overlook. Studies have reported that MYCaW® effectively identifies key areas of distress among cancer patients, facilitating tailored interventions that improve overall quality of life [10]. For example, MYCaW® has been used in palliative care settings to evaluate holistic treatment approaches, where its open-ended nature provided valuable qualitative insights into patient needs [11]. Here, the MYCaW® questionnaire was well-accepted by healthcare professionals working with frail individuals, with most finding it easy to use and beneficial for patient-focused discussions. A standardized coding framework with five super-categories and 36 specific categories was developed to analyze concerns, achieving high inter-rater reliability (κ = 0.905) [11]. Additionally, studies comparing MYCaW® with traditional HRQL measures, such as the EORTC QLQ-C30, have found that MYCaW® complements existing tools by offering a more individualized assessment [12].

Like MYCaW®, the precursor questionnaire *Measure Yourself Medical Outcome Profile* (MYMOP®) has also been used extensively in clinical settings to evaluate patient-centered outcomes. While MYMOP® is more focused on symptom-specific concerns and treatment effects, both instruments share a common philosophy of prioritizing patient voice and individualized health concerns [13]. Studies have highlighted that MYMOP® and MYCaW® provide complementary insights, with MYCaW® being particularly advantageous in psycho-oncology due to its focus on well-being.

Regarding translation and cultural adaptation, previous efforts have successfully adapted MYCaW® into seven different languages, such as Danish, Polish, Hebrew, Romanian, Somali, Spanish, and Turkish following rigorous methodologies. The translation and cultural adaptation of MYCaW® into German, as outlined in the iSWOP study protocol, follows internationally recognized frameworks, including ISPOR and COSMIN guidelines [6] and meets the Meaningful Measures protocol standards. The systematic translation process ensures that the adapted version maintains conceptual equivalence and cultural relevance [5].

Past translation efforts of PROMs have underscored the importance of meticulous linguistic validation to preserve the tool’s validity and reliability. The German MYCaW® adaptation aligns with similar initiatives in other languages, which have emphasized the need for iterative refinement through patient and expert feedback [14]. These processes are essential for ensuring that the instrument effectively captures the lived experiences of diverse patient populations.

Despite its strengths, the adaptation of MYCaW® into German also presents challenges. The preliminary validation sample size, while adequate for initial testing, may not fully capture the heterogeneity of the German-speaking cancer population. Furthermore, potential biases in translation, particularly in conveying nuanced emotional and psychological concerns, could affect the instrument’s equivalence across languages. Future studies should focus on larger-scale validation and psychometric testing to further refine the German MYCaW® and enhance its applicability in routine clinical practice [15].

In conclusion, MYCaW® has proven to be a valuable tool for assessing individualized patient concerns and well-being in oncology care. Its successful application in various studies highlights its relevance, while the ongoing translation and adaptation efforts aim to expand its accessibility across different linguistic and cultural contexts. The German MYCaW® version, once validated, will provide an essential resource for patient-centered research and care in the German-speaking populations, ultimately contributing to a more comprehensive understanding of cancer survivors’ experiences and needs.

### Trial status

The current protocol version is 1.0, dated from 17.01.2023. The trial is ongoing and currently enrolling. The first participant was enrolled on 23.01.2023 and recruitment is expected to be completed in the 2nd quarter of 2025 with the follow-up completed by the end of 2025.

## Data Availability

All data produced in the present work are contained in the manuscript.

## Footnotes

### Patient and public involvement

Patients and/or the public were not involved in the design, or conduct, or reporting, or dissemination plans of this research.

## Acknowledgements

We would like to thank Antje Merkle, Stephan Popp, Irena Cop, Danilo Pranga, and Andreas Matthes at the Havelhöhe Hospital for their support in this work.

## Ethics approval and consent to participate

This study adheres to the principles of the Declaration of Helsinki and has received approval from the Ethics Committee of the Berlin Medical Association (Ethik-Kommission der Ärztekammer Berlin) under reference number Eth-27/10. This trial is registered in the German Clinical Trials Register (DRKS00013335, registered on 27/11/2017). Written informed consent will be obtained from all participants before enrollment. Any significant protocol deviations or modifications will be reported to the Ethics Committee and the German Clinical Trials Register.

## Availability of data and materials

All data produced in the present work are contained in the manuscript.

## Consent for publication

Model consent forms will be provided on request.

## Authors’ contributions

AT conceived the study, initiated the study design, developed the methodology, translated the questionnaire from English to German (forward translation), wrote the first draft of the manuscript and reviewed it, and is responsible for data curation, formal analysis, validation, and visualization, KA translated the questionnaire from German to English (backward translation). LS, CJ, SLO, SJ, JR conceived the study, and reviewed the manuscript. FS conceived the study, initiated the study design, developed the methodology, translated the questionnaire from English to German (forward translation), and is responsible for funding acquisition, validation, visualization, writing, reviewing of the manuscript, and project administration.

